# Sexual exposures associated with mpox infection: California, November 2022 to June 2023

**DOI:** 10.1101/2023.08.07.23293793

**Authors:** Robert E. Snyder, Kayla Saadeh, Eric C. Tang, Kelly A. Johnson, Samuel N. Holland, Joshua Quint, Nicole O. Burghardt, Shua J. Chai, Roshani Fernando, Kimberly Gonzalez Barrera, Cindy Hernandez, Korie McManus, Kieran Lorenz, Jarett Maycott, John McGinley, Joseph A. Lewnard

**Affiliations:** California Department of Public Health, Richmond and Sacramento, CA, USA; School of Public Health, University of California, Berkeley, CA, USA; Career Epidemiology Field Officer Program, Division of State and Local Readiness, Office of Readiness and Response, Centers for Disease Control and Prevention, Atlanta, GA, USA

## Abstract

**Background:** Exposures associated with mpox infection remain imperfectly understood.

**Methods:** We conducted a case-control study enrolling participants who received molecular tests for mpox/orthopoxvirus in California from November 2022 through June 2023. We collected data on behaviors during a 21-day risk period before symptom onset or testing among mpox cases and test-negative controls.

**Results:** Thirteen of 54 (24.1%) cases and 5/117 (4.3%) controls reported sexual exposure to individuals they identified as potential mpox cases (“index contacts”; odds ratio [OR]=7.7 [95% confidence interval: 2.5-19.3]). Among these participants, 10/13 (76.9%) cases and 2/5 (40.0%) controls reported their index contacts were not experiencing symptoms visible to participants during sex (OR=14.9 [3.6-101.8]). Only 3/54 cases (5.6%) recalled exposure to symptomatic index contacts. Cases reported greater numbers of anal/vaginal sex partners than controls (adjusted OR=2.2 [1.0-4.8] for 2-3 partners and 3.8 [1.7-8.8] for ≥4 partners). Male cases with penile lesions more commonly reported insertive anal/vaginal sex than those without penile lesions (adjusted OR=9.3 [1.6-54.8]). Cases with anorectal lesions more commonly reported receptive anal sex than cases without anorectal lesions (adjusted OR=14.4 [1.0-207.3]).

**Conclusions:** Sexual exposure to contacts known or suspected to have experienced mpox was associated with increased risk of infection, often when index contacts lacked apparent symptoms. Exposure to greater numbers of sex partners, including those whom participants did not identify as index contacts, was associated with increased risk of infection in a site-specific manner. While participants’ assessment of symptoms in partners may be imperfect, these findings suggest individuals without visibly prominent mpox symptoms transmit infection.

## INTRODUCTION

An ongoing, multi-country mpox outbreak has involved sustained human-to-human spread of human mpox/monkeypox virus (hMPXV) concentrated among men who have sex with men (MSM). Current guidance by the US Centers for Disease Control and Prevention (CDC) emphasizes close, skin-to-skin contact with rashes or scabs from infected people as a key exposure driving transmission, stating that people with mpox can spread infection to others from the time symptoms start until a rash has fully healed and a fresh layer of skin has formed.^1^ This guidance also acknowledges the possibility of presymptomatic transmission 1-4 days before symptom onset, but states that there is no evidence that people who never develop symptoms can spread hMPXV to someone else.

Among people infected with hMPXV, detection of viral genetic material has been reported from a variety of specimens including lesion-unaffected skin, saliva, urine, blood, semen, feces, and on swabs taken from the oropharynx, anus, rectum, and genitals.^2–6^ Additionally, replication-competent hMPXV has been isolated from anorectal swabs of cases who remained asymptomatic throughout their infection.^7^ While presymptomatic hMPXV transmission has been identified in anecdotal reports,^8,9^ the contribution of visible lesions to transmission by infected individuals has not been established.

Understanding hMPXV transmission is of key importance to public health efforts aimed at identifying individuals at risk for infection^10,11^ and communicating effective risk-management strategies.^12^ We undertook a test-negative design case-control study aiming to identify risk factors for infection among individuals who received mpox testing in California from November 2022 through June 2023.

## METHODS

### Design

Clinical providers and laboratories in California report all hMPXV or orthopoxvirus tests to the California Department of Public Health (CDPH). We defined cases as individuals with positive results from any hMPXV/orthopoxvirus test (Council of State and Territorial Epidemiologists case definition) reported to CDPH during the study period. Controls were individuals who tested negative for hMPXV/orthopoxvirus without an accompanying positive or indeterminate test result (e.g., if multiple lesions were sampled from a single patient). We called cases and controls by telephone, making up to five call or voicemail message attempts per enrollee. Eligible participants were ≥18 years old, were tested 14-30 days prior to the interview, and spoke sufficient English or Spanish to provide informed consent to participate by telephone. At the request of the Los Angeles County Department of Public Health, Los Angeles County residents (except those residing in Pasadena or Long Beach) were not invited to participate. Cases in Los Angeles closely resembled those in the rest of the state in terms of gender (96% men, 2% women, 2% transgender, non-binary, or other across both jurisdictions), race/ethnicity (45-48% Hispanic/Latino, 25-31% White, 13-17% Black, 4-6% Asian across both jurisdictions), and age distribution.^13,14^ The study protocol received a non-research determination from the Committee for the Protection of Human Subjects of the California Health and Human Services Agency.

### Exposures

We used a standardized, computer-guided interview form (Qualtrics; Provo, Utah) to collect data from participants. Interview items addressed participants’ demographic and clinical characteristics (including age, sex assigned at birth [e.g., male, female] and gender identity [e.g., man, woman, transgender, non-binary], race/ethnicity, and symptoms compatible with mpox), sexual behaviors with all partners during the 21 days preceding dates of symptom onset or testing (whichever was earliest; “risk period”), and whether participants were aware of any interaction with a potential mpox case (defined as a “potential index contact” here and below) during this risk period. Questions on sexual encounters with individuals not identified as potential index contacts were introduced in December after enrollment had begun. Using first name, last name, and date of birth, we cross-referenced participants against the CDPH Office of AIDS HIV case registry to determine HIV infection status, with the CDPH STI case registry to determine chlamydia, gonorrhea, and syphilis infection history, and with the California Immunization Registry to determine JYNNEOS vaccination status.

We requested that participants recall any interactions with (1) individuals who had symptoms of mpox during their interaction, or (2) individuals who participants learned may have had mpox after the interaction occurred. Participants who answered “yes” to either type of exposure were asked to specify if they were aware that their contacts were diagnosed with mpox by a healthcare provider (“diagnosed index contacts”) or if they were unaware whether their contacts received any such diagnosis (“suspect index cases”). To ensure capture of any relevant symptoms in index contacts, questions about index contact exposures followed an earlier questionnaire block addressing participants’ own experience with all potential mpox symptoms listed on the CDC case report form;^15^ interviewers maintained a list of these symptoms for clarification, if needed, during calls. For participants who reported exposure to a diagnosed or suspect index contact, we asked whether this exposure involved long-lasting face-to-face contact (≥3 hours); touching one another’s skin; touching shared fomites (e.g., food/dishes/utensils, towels/bedding/clothing; or drugs/drug equipment); providing care to the index case while they were sick; or sexual contact (e.g., intimate touching, use of shared sex toys, oral sex, or anal/vaginal intercourse).

As this study was not a component of public health contact tracing activities, we did not collect identifying information on potential index contacts from participants and could not verify whether they had been tested or had received positive results.

### Statistical analysis

We first aimed to determine the association of hMPXV infection status with participants’ knowledge of exposure to a potential diagnosed or suspected index contact within the 21-day risk period. We computed odds ratios (ORs) and accompanying 95% confidence intervals (CIs) comparing the odds of recall of the following exposures between cases and controls: any exposure to potential index contacts; non-sexual exposure to potential index contacts; sexual exposure to potential index contacts; sexual exposure to potential index contacts whom participants recalled as experiencing symptoms; and sexual exposure to potential index contacts whom participants did not recall as experiencing symptoms. We analyzed each of these exposures separately for diagnosed and suspect index contacts. All analyses defined no known contact (sexual or non-sexual) with a potential index contact as the reference exposure.

Because participants with greater numbers of sex partners may have been more likely to believe, by chance, that one could have been an mpox case, we stratified analyses by participants who reported intimate touching, oral sex, anal/vaginal sex, or use of sex toys with multiple partners during the risk period and those who did not report multiple sexual partners. Last, because HIV infection could modify the likelihood of hMPXV infection, given exposure, we also stratified these analyses by participants’ HIV infection status.

Because few participants reported exposure to potential index contacts, we also aimed to identify whether sexual partnerships not known to involve index contacts were associated with risk of infection. To mitigate potential confounding driven by differences in risk status between cases and controls, or the willingness of cases and controls to report sexual exposures, we restricted these analyses to cases and controls who provided information on ≥1 sexual partnership during the risk period. We computed ORs measuring the association of case or control status with the number of partners with whom participants reported any intimate touching, any oral sex (giving or receiving), any anal or vaginal sex (insertive or receptive), and any anal or vaginal sex without condoms. We computed adjusted ORs for each exposure using conditional logistic regression models matching participants on sex and whether they reported contact with any potential index contact. Analyses addressing each sexual exposure adjusted for the number of partners with whom participants reported engaging in all other sexual acts listed above.

To assess the biological plausibility that reported exposures accounted for hMPXV infection among cases, we further estimated associations of specific reported sex acts with sites of lesion occurrence among cases who reported lesions. We compared odds of intimate touching of the penis, receptive oral sex involving the penis, and insertive anal or vaginal sex (with or without condoms) among male cases reporting penile lesions or no penile lesions during their illness.

Additionally, we compared odds of intimate touching of the anus/rectum, receptive oral sex on the anus/rectum, and receptive anal sex (with or without condoms) among cases (male or female) reporting anorectal lesions or no anorectal lesions during their illness. For intimate touching and oral sex exposures, we repeated these analyses within subgroups limited to participants who did not report condomless anal or vaginal sex acts. Because we enrolled few female cases, we did not undertake similar analyses comparing cases with or without vaginal lesions. We used logistic regression to adjust for other sex acts reported by participants.

We conducted statistical analyses in R (version 4.3.0; R Foundation for Statistical Computing, Vienna, Austria).

## RESULTS

### Enrollment

Between November 2022 and June 2023, we enrolled 54 mpox cases and 117 controls (**Table 1**). In total, 49 (90.7%) cases and 92 (78.6%) controls were assigned male sex at birth, and 48 (88.9%) cases and 92 (78.6%) controls were cisgender men. We also enrolled 2 transgender man and 1 transgender woman as cases. Among 49 male cases, 39 (88.9%) reported male sex partners during the risk period, versus 32/92 (34.8%) male controls (64.0% of 50 male controls who provided information on ≥1 sex partner during the risk period). Racial and ethnic composition was similar among cases and controls. Cases were more likely than controls to have HIV infection, to have laboratory-confirmed history of chlamydia, gonorrhea, or syphilis, and to have received ≥1 JYNNEOS vaccine dose. Differences in vaccination among cases and controls were mitigated within analyses subset to MSM and non-MSM strata, in accordance with prioritization tiers under the US National Mpox Vaccination Strategy^1^ (**Table S1**).

**Table 1:** Characteristics of enrolled cases and test-negative controls.

| Characteristic |  | Prevalence, *n* (%) |  |
| --- | --- | --- | --- |
|  |  | Cases | Test-negative controls |
|  |  | *N*=54 | *N*=117 |
| Sex |  |  |  |
|  | Male at birth | 49 (90.7) | 92 (78.6) |
|  | Female at birth | 5 (9.3) | 25 (21.4) |
| Gender |  |  |  |
|  | Cisgender men | 48 (88.9) | 92 (78.6) |
|  | Cisgender women | 3 (5.6) | 25 (21.4) |
|  | Transgender men | 2 (3.7) | 0 |
|  | Transgender women | 1 (1.9) | 0 |
| Sexual behavior during risk period (among participants assigned male sex at birth) |  |  |  |
|  | Male sex partners during risk period | 39/49 (79.6) | 32/92 (34.8) |
|  | No male sex partners or no recent sex partners identified during risk period<sup>1</sup> | 10/49 (20.4) | 60/92 (65.2) |
| Race or ethnicity<sup>1</sup> |  |  |  |
|  | White | 24 (44.4) | 67 (57.3) |
|  | Hispanic | 23 (42.6) | 48 (41.0) |
|  | Asian | 5 (9.3) | 10 (8.5) |
|  | Black or African American | 5 (9.3) | 6 (5.1) |
|  | Native Hawaiian or Pacific Islander | 1 (1.9) | 2 (1.7) |
|  | American Indian or Alaska Native | 1 (1.9) | 3 (2.6) |
| Self-reported history of chlamydia, gonorrhea, and syphilis |  |  |  |
|  | No history of infection reported | 18 (33.3) | 75 (64.1) |
|  | Infection <3 weeks previously | 10 (18.5) | 5 (4.3) |
|  | Infection ≥3 weeks and <12 months previously | 8 (14.8) | 13 (11.1) |
|  | Infection ≥1 year previously | 18 (33.3) | 24 (20.5) |
| Self-reported history of other sexually transmitted infections |  |  |  |
|  | No history of infection reported | 49 (90.7) | 95 (81.2) |
|  | Infection <3 weeks previously | 0 | 7 (6.0) |
|  | Infection ≥3 weeks and <12 months previously | 0 | 0 |
|  | Infection ≥1 year previously | 5 (9.3) | 15 (12.8) |
| HIV infection<sup>2</sup> |  |  |  |
|  | Living with HIV infection | 19 (35.2) | 18 (15.4) |
|  | Not living with HIV infection | 35 (64.8) | 99 (84.6) |
| History of JYNNEOS vaccination |  |  |  |
|  | No vaccination | 34 (63.0) | 89 (76.1) |
|  | 1 vaccine dose | 8 (14.8) | 12 (10.3) |
|  | 2 vaccine doses | 12 (22.2) | 16 (13.7) |

<sup>1</sup>Calculated among male participants who provided information on ≥1 sex partner during the risk period, 39/44 (88.9%) of cases and 32/50 (64.0%) of controls had male sex partners.
<sup>2</sup>We indicate all races or ethnicities listed by participants; totals do not sum to 100% as individuals could report identifying with multiple racial or ethnic categories.
<sup>3</sup>We verified HIV infection status against CDPH Office of AIDS HIV case registry data.

### Exposure to potential index mpox contacts

Seventeen (30.8%) cases and 7 (6.0%) controls reported exposure to a potential index contact during the risk period (OR=7.2 [95% CI: 2.5-13.5] among cases versus controls; **Table 2**). Eight cases and 4 controls identified their potential index contact as a diagnosed index contact (OR=5.9 [95% CI: 1.7-19.9] among cases versus controls). Most cases (13/17; 76.5%) and controls (5/7; 71.4%) indicated that their exposures to potential index contacts were sexual (OR=7.7 [2.5-19.3] among cases versus controls). Point estimates also indicated higher odds of non-sexual exposure to potential index contacts among cases than controls (OR=5.9 [1.1-47.3] among cases vs. controls).

**Table 2:** Recall of exposure to mpox index contacts among participants.

| Exposure |  | Prevalence of exposure, *n*<br>(%) |  | OR (95% CI)<sup>†</sup> |
| --- | --- | --- | --- | --- |
|  |  | Cases | Test-negative controls |  |
|  |  | *N*=54 | *N*=117 |  |
| Exposure to an index contact | Any index contact | 17 (31.5) | 7 (6.0) | 7.2 (2.5, 13.5) |
|  | Diagnosed index contact | 8 (14.9) | 4 (3.4) | 5.9 (1.7, 19.9) |
|  | Suspect index contact | 9 (16.7) | 3 (2.6) | 8.9 (2.4, 37.0) |
| Non-sexual exposure to an index contact | Any index contact | 4 (7.4) | 2 (1.7) | 5.9 (1.1, 47.3) |
|  | Diagnosed index contact | 2 (3.7) | 2 (1.7) | 3.0 (0.3, 27.3) |
|  | Suspect index contact | 2 (3.7) | 0 | — |
| Sexual exposure to an index contact | Any index contact | 13 (24.1) | 5 (4.3) | 7.7 (2.5, 19.3) |
|  | Diagnosed index contact | 6 (11.1) | 2 (1.7) | 8.9 (2.0, 66.3) |
|  | Suspect index contact | 7 (13.0) | 3 (2.6) | 6.9 (1.8, 30.6) |
| Sexual exposure to an index contact with apparent symptoms at the time of the encounter | Any index contact | 3 (5.6) | 3 (2.6) | 3.0 (0.5, 15.9) |
|  | Diagnosed index contact | 1 (1.9) | 1 (0.9) | 3.0 (0.1, 111.3) |
|  | Suspect index contact | 2 (3.7) | 2 (1.7) | 3.0 (0.3, 27.3) |
| Sexual exposure to an index contact without apparent symptoms at the time of the encounter | Any index contact | 10 (18.5) | 2 (1.7) | 14.9 (3.6, 101.8) |
|  | Diagnosed index contact | 5 (9.3) | 1 (0.9) | 14.9 (2.5, 531.8) |
|  | Suspect index contact | 5 (9.3) | 1 (0.9) | 14.9 (2.5, 531.8) |

Abbreviations: OR—odds ratio; CI: confidence interval.
We define “exposure to an index contact” as any recollection of a partner experiencing mpox symptoms during an interaction, or recollection of interaction with a partner who participants subsequently learned may have had mpox. Participants were asked to specify whether they were aware of index cases having received an mpox diagnosis from a healthcare provider, regardless of their answer to the previous questions.
<sup>†</sup>All analyses define individuals without any recall of exposure to an index case as the referent group (37 cases [68.5% of 54]; 110 controls [94.0% of 117]).

Among 13 cases and 5 controls reporting sexual exposure to potential index contacts, 10 cases and 2 controls could not recall their contact experiencing visibly apparent symptoms during encounters (OR=14.9 [3.6-101.8] for cases versus controls; **Table 2**). This association was apparent for exposures to diagnosed as well as suspect index contacts. We obtained similar findings in analyses stratified for participants who reported or did not report multiple partnerships during the study period (**Table S2**; **Table S3**). Within the full sample, only 3 cases (5.6%) and 3 controls (2.6%) reported sexual exposure to potential index contacts whom participants could recall as experiencing mpox symptoms at the time of the encounter (OR=3.0 [0.5-15.9]; **Table 2**).

Seven of 19 cases with HIV infection (36.8%) reported exposure to potential index contacts, among whom 6 (31.6%) reported sexual exposure to a potential index case (**Table 3**). No controls with HIV infection reported exposure to potential index contacts, precluding numerical analyses within this subgroup. Among HIV-uninfected participants, cases had higher odds than controls of reporting any exposure to a potential index contact (OR=5.3 [1.7-10.7] among cases versus controls) and sexual exposure to a potential index contact (OR=5.2 [1.5-14.4] among cases versus controls).

**Table 3:**
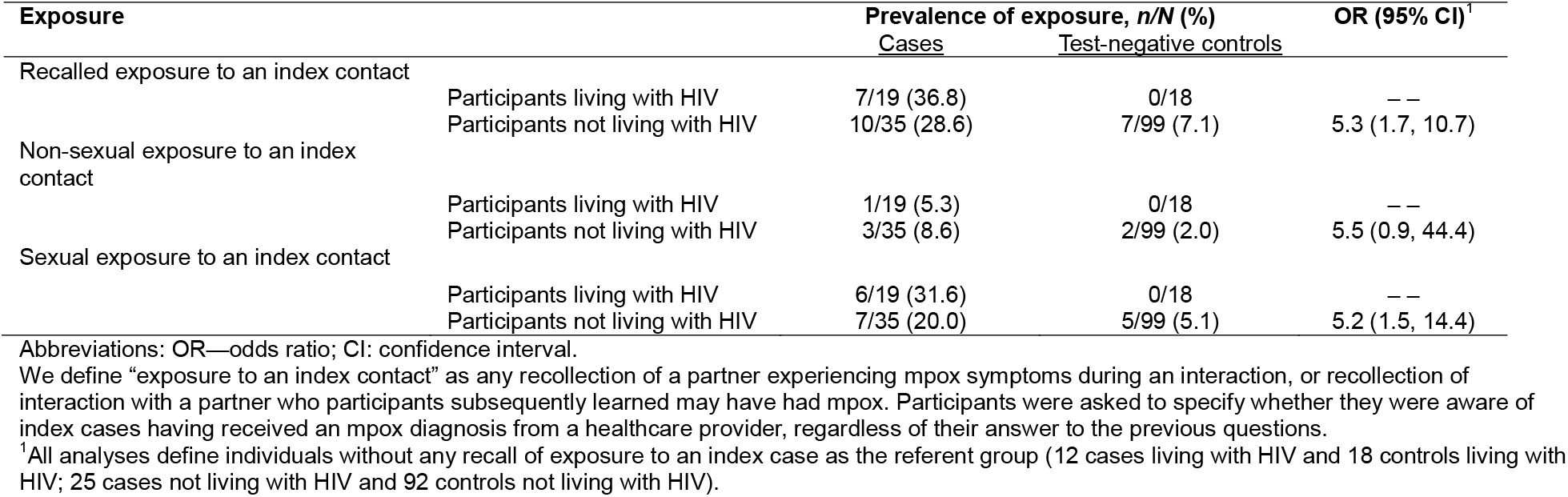
Recall of exposure to mpox index contacts among participants with and those without HIV infection.

### All sexual exposures among cases and controls

Information on sexual acts undertaken with ≥1 partner during the risk period were available from 37 cases and 65 controls (**Table 4**). Comparing cases to controls, unadjusted odds ratios of reporting the behaviors listed below with 2-3 or ≥4 partners (versus 0-1 partners) were 3.4 (1.3-9.0) and 3.9 (1.5-10.3), respectively, for intimate touching; 1.5 (0.8-2.7) and 2.2 (1.2-4.0), respectively, for oral sex; 2.5 (1.2-5.0) and 3.6 (1.8-7.1), respectively, for anal/vaginal sex; and 2.1 (1.2-3.7) and 2.7 (1.5-4.9), respectively, for condomless anal/vaginal sex. After adjustment for reporting of multiple sex acts and exposure to potential index cases, however, statistically significant evidence of associations (two-sided *p*<0.05) persisted for only some exposures. Comparing cases to controls, adjusted odds ratios of reporting anal/vaginal sex with 2-3 and ≥4 partners (versus 0-1 partners) were 2.2 (1.0-4.8) and 3.8 (1.7-8.8), respectively. For anal/vaginal sex without condoms, corresponding adjusted odds ratios were 2.3 (1.2-4.4) and 3.6 (1.5-8.8), respectively. In addition, cases had 3.0 (1.0-9.0) and 2.9 (0.9-9.4) fold higher adjusted odds of reporting intimate touching with 2-3 and ≥4 partners (versus 0-1 partners) in comparison to controls.

**Table 4:** Sexual exposures among participants providing information on ≥1 sexual encounter during the risk period.

| Exposure |  | Prevalence of exposure, n/N (%) |  | OR (95% CI) |  |
| --- | --- | --- | --- | --- | --- |
|  |  | Cases | Test-negative controls | Unadjusted | Adjusted<sup>1</sup> |
|  |  | N=37 | N=65 |  |  |
| Intimate touching | Participants reporting any exposure | 37 (100.0) | 62 (95.4) | -- | -- |
|  | Participants reporting 0-1 partners | 4 (10.8) | 27 (41.5) | ref. | ref. |
|  | Participants reporting 2-3 partners | 16 (43.2) | 21 (32.3) | 3.4 (1.3, 9.0) | 3.0 (1.0, 9.0) |
|  | Participants reporting ≥4 partners | 17 (45.9) | 17 (26.2) | 3.9 (1.5, 10.3) | 2.9 (0.9, 9.4) |
| Oral sex | Participants reporting any exposure | 33 (89.2) | 58 (89.2) | -- | -- |
|  | Participants reporting 0-1 partners | 14 (37.8) | 38 (58.5) | ref. | ref. |
|  | Participants reporting 2-3 partners | 13 (35.1) | 20 (30.8) | 1.5 (0.8, 2.7) | 0.9 (0.4, 1.8) |
|  | Participants reporting ≥4 partners | 10 (27.0) | 7 (10.8) | 2.2 (1.2, 4.0) | 0.7 (0.3, 1.8) |
| Anal/vaginal sex | Participants reporting any exposure | 36 (97.3) | 58 (89.2) | -- | -- |
|  | Participants reporting 0-1 partners | 9 (24.3) | 40 (61.5) | ref. | ref. |
|  | Participants reporting 2-3 partners | 16 (43.2) | 19 (29.2) | 2.5 (1.2, 5.0) | 2.2 (1.0, 4.8) |
|  | Participants reporting ≥4 partners | 12 (32.4) | 6 (9.2) | 3.6 (1.8, 7.1) | 3.8 (1.7, 8.8) |
| Anal/vaginal sex without condoms | Participants reporting any exposure | 31 (83.8) | 54 (83.1) | -- | -- |
|  | Participants reporting 0-1 partners | 16 (43.2) | 49 (75.4) | ref. | ref. |
|  | Participants reporting 2-3 partners | 13 (35.1) | 12 (18.5) | 2.1 (1.2, 3.7) | 2.3 (1.2, 4.4) |
|  | Participants reporting ≥4 partners | 8 (21.6) | 4 (6.2) | 2.7 (1.5, 4.9) | 3.6 (1.5, 8.8) |
| Anal/vaginal sex with condoms<sup>2</sup> | Participants reporting any exposure | 14 (37.8) | 15 (23.1) | -- | -- |
|  | Participants reporting 0-1 partners | 31 (83.8) | 59 (90.8) | ref. | ref. |
|  | Participants reporting 2-3 partners | 5 (13.5) | 4 (6.2) | 1.6 (0.8, 3.1) | 1.6 (0.8, 3.1) |
|  | Participants reporting ≥4 partners | 1 (2.7) | 2 (3.1) | 1.0 (0.2, 4.9) | 1.2 (0.2, 10.0) |

Abbreviations: OR—odds ratio; CI: confidence interval.
We estimate adjusted associations in logistic regression models accounting for participant sex, recall of exposure to a potential index case, and other reported sexual acts performed; analyses adjust for the number of partners with whom participants report engaging in all listed behaviors, and match on participants' history of sexual contact with a potential index case and sex (male/female).

Male cases with penile lesions had 9.3 (1.6-54.8) fold higher adjusted odds of reporting insertive anal/vaginal sex acts with any partner during the risk period in comparison to male cases who did not experience penile lesions (**Table 5**). Likewise, cases with anorectal lesions had 14.4 (1.0-207.3) fold higher adjusted odds of reporting receptive anal sex with any partner during the risk period than cases without anorectal lesions. We did not identify strong evidence that intimate touching or oral sex involving the penis or anus/rectum were associated with occurrence of lesions at either site among cases. Additionally, insertive anal/vaginal sex and receptive anal sex acts with condoms were not independently associated with penile or anorectal lesion occurrence, respectively (adjusted OR=1.6 [0.1-25.5] and 0.9 [0.2-5.2], respectively), although few participants reported condom use during anal or vaginal sex.

**Table 5:**
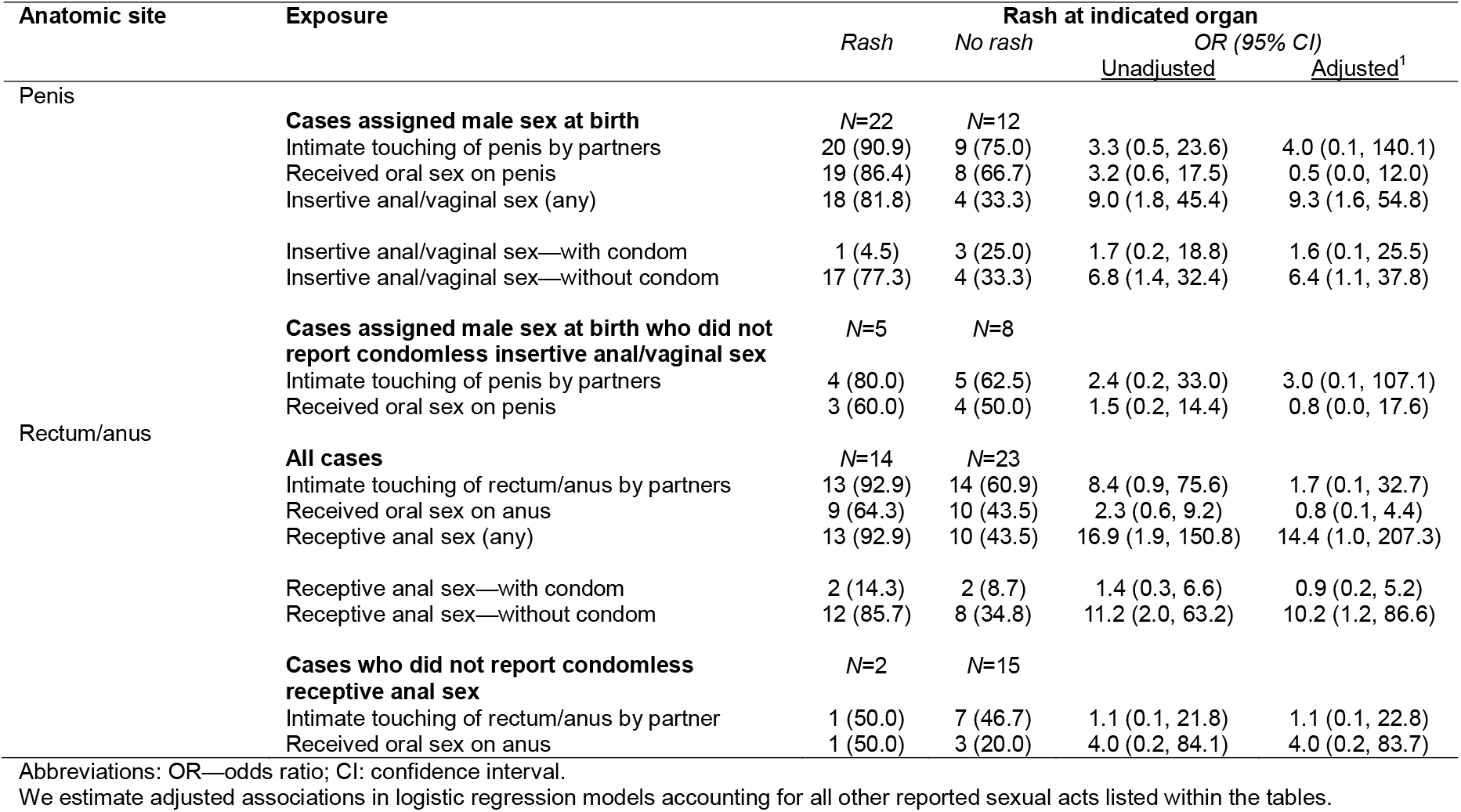
Rash location and site-specific sexual exposures among cases.

In stratified analyses, odds of reporting intimate touching or oral sex exposures on the penis among HIV-negative cases were 18.0 (1.3-702.9) and 14.8 (1.5-245.5) fold higher, respectively, among cases with penile lesions than cases without penile lesions (**Table 6**). Among HIV-negative cases who did not report penile lesions, none reported insertive anal/vaginal sex acts. Odds of reporting intimate touching and receptive oral sex on the anus/rectum were 6.4 (0.7-207.5) and 14.5 (1.5-473.1) fold higher, respectively, among HIV-negative cases who had anorectal lesions as compared to those without anorectal lesions. All HIV-negative cases who experienced anorectal lesions reported receptive anal sex, and 6/7 reported condomless receptive anal sex (OR=19.5 [2.0-650.4]).

**Table 6:** Rash location and site-specific sexual exposures among cases with and those without HIV.

| Anatomic site | Exposure | Rash | No rash | OR (95% CI)<br>Unadjusted |
| --- | --- | --- | --- | --- |
| Penis (male cases only) | **Cases living with HIV and assigned male sex at birth** | N=10 | N=6 |  |
|  | Intimate touching of penis by partners | 8 (80.0) | 4 (66.7) | 2.1 (0.2, 28.9) |
|  | Received oral sex on penis | 8 (80.0) | 4 (66.7) | 2.1 (0.2, 28.9) |
|  | Insertive anal/vaginal sex (any) | 8 (80.0) | 3 (50.0) | 4.7 (0.5, 65.0) |
|  | Insertive anal/vaginal sex—with condom | 0 | 0 | — |
|  | Insertive anal/vaginal sex—without condom | 8 (80.0) | 3 (50.0) | 4.7 (0.5, 65.0) |
|  | **Cases not living with HIV and assigned male sex at birth** | N=13 | N=6 |  |
|  | Intimate touching of penis by partners | 12 (92.3) | 3 (50.0) | 18.0 (1.3, 702.9) |
|  | Received oral sex on penis | 11 (84.6) | 2 (33.3) | 14.8 (1.5, 245.5) |
|  | Insertive anal/vaginal sex (any) | 11 (84.6) | 0 | — |
|  | Insertive anal/vaginal sex—with condom | 2 (15.4) | 0 | — |
|  | Insertive anal/vaginal sex—without condom | 9 (69.2) | 0 | — |
| Rectum/anus | **Cases living with HIV** | N=7 | N=11 |  |
|  | Intimate touching of rectum/anus by partners | 5 (71.4) | 6 (54.5) | 2.3 (0.3, 25.0) |
|  | Received oral sex on anus | 2 (28.6) | 5 (45.5) | 0.4 (0.0, 3.4) |
|  | Receptive anal sex (any) | 5 (71.4) | 5 (45.5) | 3.4 (0.4, 37.8) |
|  | Receptive anal sex—with condom | 1 (14.3) | 1 (9.1) | 1.7 (0.0, 76.7) |
|  | Receptive anal sex—without condom | 4 (57.1) | 4 (36.4) | 2.5 (0.3, 21.8) |
|  | **Cases not living with HIV** | N=7 | N=16 |  |
|  | Intimate touching of rectum/anus by partners | 6 (85.7) | 9 (56.2) | 6.4 (0.7, 207.5) |
|  | Received oral sex on anus | 6 (85.7) | 6 (37.5) | 14.5 (1.5, 473.1) |
|  | Receptive anal sex (any) | 7 (100.0) | 9 (56.2) | — |
|  | Receptive anal sex—with condom | 3 (42.9) | 4 (25.0) | 2.3 (0.3, 17.8) |
|  | Receptive anal sex—without condom | 6 (85.7) | 5 (31.2) | 19.5 (2.0, 650.4) |

Abbreviations: OR—odds ratio; CI: confidence interval.

## DISCUSSION

Our analyses provide evidence suggesting that individuals without visibly prominent symptoms of mpox may transmit hMPXV infection. First, few cases enrolled in our study recalled sexual exposure to potential index contacts experiencing symptoms that were apparent to participants at the time of the encounter. A greater proportion recalled exposure to potential index contacts who were not experiencing apparent symptoms, and these exposures were associated with increased risk of infection. Additionally, anal or vaginal intercourse with greater numbers of partners was independently associated with increased risk of infection after adjusting for participants’ known exposure to potential index contacts, supporting the hypothesis that exposures to partners not identified as potential index contacts accounted for a substantial share of hMPXV acquisition in our sample. Moreover, recent insertive and receptive sex acts were associated with lesion occurrence at penile and anorectal anatomic sites, respectively. Collectively, the low proportion of cases recalling exposure to symptomatic individuals, the association of infection risk with sexual encounters involving individuals who did not experience mpox symptoms, and the specificity of this association by site of sexual exposure and lesion occurrence suggest hMPXV can be acquired through sexual contact with individuals not experiencing prominent mpox symptoms.

Whereas current US public health guidance emphasizes close, skin-to-skin contact with rashes or scabs from cases with mpox as the primary route of transmission for hMPXV,^1^ our analyses complement other recent data that suggest the potential for transmission by individuals without symptoms.^16^ Contact-tracing studies in the UK^8^ and the Netherlands,^9^ that established links between confirmed cases identified pre-symptomatic transmission in 28-80% of all confirmed case pairs. However, such studies may be prone to underestimating the frequency of transmission by persons without symptoms due to the potential for cases to go undiagnosed or for partnerships to go unascertained. This risk may be especially pronounced in the context of anonymous encounters. Within our study, anonymous sex may have hindered participants’ ability to identify instances where they were exposed to presymptomatic individuals with whom they had no further contact, including after these individuals experienced symptoms. For this reason, our finding that 10 of 54 cases reported contact with potential index cases while they were not experiencing symptoms may underestimate the frequency of pre-symptomatic transmission. However, anonymous sex is not likely to account for the low proportion of cases who recalled exposure to partners while these partners were experiencing apparent symptoms (3 of 54 cases). This observation suggests that exposure to symptomatic individuals may account for a smaller-than-expected proportion of mpox cases.

Three recent studies in which MSM were prospectively tested for hMPXV DNA via anorectal swabs found that a majority of detections were associated with infections that did not ultimately result in symptoms or care-seeking;^7,17,18^ in one study, these asymptomatic infections were confirmed to result in seroconversion and replication-competent viral shedding.^7^ Many patients report significant pain and discomfort associated with mpox lesions,^19–21^ which may reduce their likelihood of pursuing sexual contact while lesions are present. While US public health guidance indicates that no cases of transmission have been definitively linked to exposure to infected persons who never developed signs or symptoms of illness (i.e., asymptomatic infection), opportunities to identify such infections are limited by guidelines allowing only for lesion-based diagnostic testing.^1^ This may lead to missed opportunities for identifying individuals at risk of spreading hMPXV, as well as inaccurate estimates of the incidence and prevalence of hMPXV infection.^2–6^

In our study, all participants with HIV infection who reported exposure to a potential index case were infected with mpox, precluding direct comparisons of the association of infection risk with participants’ recall of exposure to index cases among participants according to HIV infection status. Attenuated effect size estimates for this association among HIV-negative participants suggests HIV infection may be associated with the risk of acquiring infection given exposure to an mpox case. Lesion sites were also less strongly associated with sexual exposure sites among cases with HIV infection, consistent with the hypothesis that HIV enhances clinical severity of mpox. While prevalence of HIV infection was greater among cases than controls in our analysis, our analyses were underpowered to generate adjusted estimates of the effect of HIV infection on risk of hMPXV infection accounting for differences in risk of exposure among HIV-positive and HIV-negative individuals. This pattern may also be driven by effects of HIV infection on individuals’ risk of experience mpox symptoms, leading to a higher likelihood of testing and case ascertainment.

Cases reported engaging in intimate touching, oral sex, and anal/vaginal sex with greater numbers of partners than controls, suggesting intimate contact with greater numbers of partners is associated with increased risk of infection. Although we did not identify a strong association between case status and anal/vaginal sexual acts involving condoms, this finding should be interpreted cautiously. Because most reported anal/vaginal sex acts did not involve condoms, this analysis was underpowered and may not indicate protective effect modification by condom use. Similarly, the lack of an association between oral sex acts and case status may owe to correlation between the number of partnerships involving both intimate touching and oral sex.

Several limitations of our analysis should be considered. Our sample size was low due to case counts and testing effort during the study period, limiting opportunities to adjust for covariates including JYNNEOS vaccination status and HIV-related clinical variables such as viral suppression. While conducting enrollment and interviews by telephone offered an opportunity to build trust and rapport with participants, some individuals may have declined to participate due to inconvenience or concerns around disclosing personal sexual histories. These factors may introduce nonresponse bias and limit external generalizability of our findings. Whereas the population at risk for mpox includes individuals of diverse sexual orientations and gender identities, our sample was insufficient for subgroup analyses within all relevant strata of interest (e.g., cisgender MSM versus transgender participants). Last, recall of exposures over the 21-day period prior to participants’ dates of testing or symptom onset may be imperfect, including for questions around symptoms in potential index contacts. This may have led to under-estimation of the proportion of index cases who experienced symptoms at the time of sexual encounters with participants. Recall bias is also a concern in the event that cases may have been more likely to reflect on and recall potential exposures than controls, or if the test-negative control population included individuals who sought testing after very low-risk exposures.^22^ We did not ask participants whether index contacts could recall recent illnesses from which they had recovered, and some may have been in the recovery phase with lesions that were less noticeable. Regardless, public health strategies encouraging individuals to mitigate risk by avoiding partners with mpox symptoms may have limited effectiveness if symptoms are too subtle to be noticed during sexual encounters.

Our findings have several practical implications. First, the association of increased risk with greater numbers of partnerships implies that mpox incidence may be sensitive to changes in behavior. Reductions in new partnership formation among MSM during 2022 may have contributed to declining case numbers alongside vaccination, as supported by recent modeling studies,^23,24^ although avoidance of sexual contact should not be considered a viable prevention strategy in the longer term. Second, the association between lesion location and sexual practices may have relevance for interpreting clinical or surveillance data on cases, helping to identify plausible routes of exposure and potentially identify partners for testing. Third, our findings add to growing evidence that clinical symptoms may not be requisite to hMPXV transmission. While limited by our reliance on participants’ identification of potential index contacts and assessment of their symptoms, our results enhance earlier evidence of transmission by individuals without symptoms in contact-tracing studies.^8,9^ Efforts are needed to better characterize the natural history of hMPXV infection, including the risk of transmission associated with differing clinical stages and clinical presentations, particularly in settings such as California with high coverage of JYNNEOS vaccination in at-risk populations.^25^ Development of diagnostic protocols not reliant on lesion-based sampling may enhance our ability to identify individuals at risk of transmitting hMPXV.

## Data Availability

De-identified data that underlie the results reported in this article will be made available upon reasonable request to the authors.

## FOOTNOTE PAGE

### Conflicts of interest

The authors declare no competing interests.

### Sources of support

The study was supported by the CDC Strengthening STD Prevention and Control for Health Departments (STD PCHD) cooperative agreement (PS19-1901 to the California Department of Public Health STD Control Branch) and the National Institute of Allergy and Infectious Diseases (R01-AI14812701 to JAL).

### Address for correspondence

Joseph Lewnard, 2121 Berkeley Way, Room 5410, Berkeley, California 94720; Tel.: 510-664-4050.

### Data sharing statement

De-identified data that underlie the results reported in this article will be made available upon reasonable request.

### Disclaimer

The findings and conclusions in this report are those of the author(s) and do not necessarily represent the views or opinions of the California Department of Public Health, the California Health and Human Services Agency, or the US Centers for Disease Control and Prevention.

---

**Table S1:** History of STI and JYNNEOS vaccination among cases identifying or not identifying as MSM.

| Characteristic | Prevalence, *n* (%) |  |
| --- | --- | --- |
|  | Cases | Test-negative controls |
| **MSM sample** | *N*=39 | *N*=32 |
| History of chlamydia, gonorrhea, and syphilis |  |  |
| No history of infection reported | 11 (28.2) | 10 (31.2) |
| Infection within 3 weeks previously | 10 (25.6) | 3 (9.4) |
| Infection ≥3 weeks and <12 months previously | 7 (17.9) | 9 (28.1) |
| Infection ≥1 year previously | 11 (28.2) | 10 (31.2) |
| History of other sexually transmitted infections |  |  |
| No history of infection reported | 35 (89.7) | 24 (75.0) |
| Infection within 3 weeks previously | 0 | 4 (12.5) |
| Infection ≥3 weeks and <12 months previously | 0 | 0 |
| Infection ≥1 year previously | 4 (10.3) | 4 (12.5) |
| History of JYNNEOS vaccination |  |  |
| No vaccination | 22 (56.4) | 14 (43.8) |
| 1 vaccine dose received | 5 (12.8) | 8 (25.0) |
| 2 vaccine doses received | 12 (30.8) | 10 (31.2) |
| **Non-MSM sample** | *N*=15 | *N*=85 |
| History of chlamydia, gonorrhea, and syphilis |  |  |
| No history of infection reported | 7 (46.7) | 65 (76.5) |
| Infection within 3 weeks previously | 0 | 2 (2.4) |
| Infection ≥3 weeks and <12 months previously | 1 (6.7) | 4 (4.7) |
| Infection ≥1 year previously | 7 (46.7) | 14 (16.5) |
| History of other sexually transmitted infections |  |  |
| No history of infection reported | 14 (93.3) | 71 (83.5) |
| Infection within 3 weeks previously | 0 | 3 (3.5) |
| Infection ≥3 weeks and <12 months previously | 0 | 0 |
| Infection ≥1 year previously | 1 (6.7) | 11 (12.9) |
| History of JYNNEOS vaccination |  |  |
| No vaccination | 12 (80.0) | 75 (88.2) |
| 1 vaccine dose received | 3 (20.0) | 4 (4.7) |
| 2 vaccine doses received | 0 | 6 (7.1) |

**Table S2:** Recalled exposure to potential mpox index contacts among participants not reporting multiple sex partners during the risk period.

| Exposure |  | Prevalence of exposure, *n* (%) |  | OR (95% CI)<sup>1</sup> |
| --- | --- | --- | --- | --- |
|  |  | Cases | Test-negative controls |  |
|  |  | *N*=22 | *N*=41 |  |
| Exposure to an index contact | Any index contact | 7 | 2 | 9.1 (1.9, 54.9) |
|  | Diagnosed index contact | 3 | 0 | — |
|  | Suspect index contact | 4 | 2 | 5.2 (1.0, 36.7) |
| Non-sexual exposure to an index contact | Any index contact | 2 | 0 | — |
|  | Diagnosed index contact | 1 | 0 | — |
|  | Suspect index contact | 1 | 0 | — |
| Sexual exposure to an index contact | Any index contact | 5 | 2 | 6.5 (1.3, 43.0) |
|  | Diagnosed index contact | 2 | 0 | — |
|  | Suspect index contact | 3 | 2 | 3.9 (0.6, 29.7) |
| Sexual exposure to an index contact with apparent symptoms at the time of the encounter | Any index contact | 2 | 1 | 5.2 (0.5, 184.5) |
|  | Diagnosed index contact | 1 | 0 | — |
|  | Suspect index contact | 1 | 1 | 2.6 (0.1, 96.0) |
| Sexual exposure to an index contact without apparent symptoms at the time of the encounter | Any index contact | 3 | 1 | 7.8 (1.0, 257.5) |
|  | Diagnosed index contact | 1 | 0 | — |
|  | Suspect index contact | 2 | 1 | 5.2 (0.5, 184.5) |

Abbreviations: OR—odds ratio; CI: confidence interval
<sup>1</sup>All analyses define individuals without any recall of exposure to an index case as the referent group (15 cases [68.2% of 22]; 39 controls [95.1% of 41]).

**Table S3:** Recalled exposure to potential mpox index cases among participants reporting multiple sex partners during the risk period.

| Exposure |  | Prevalence of exposure, *n*<br>(%) |  | OR (95% CI)<sup>1</sup> |
| --- | --- | --- | --- | --- |
|  |  | Cases | Test-negative controls |  |
|  |  | *N*=32 | *N*=76 |  |
| Exposure to an index contact | Any index contact | 10 | 5 | 6.5 (1.9, 15.4) |
|  | Diagnosed index contact | 5 | 4 | 4.0 (1.0, 14.1) |
|  | Suspect index contact | 5 | 1 | 16.1 (2.7, 522.9) |
| Non-sexual exposure to an index contact | Any index contact | 2 | 2 | 3.2 (0.3, 28.8) |
|  | Diagnosed index contact | 1 | 2 | 1.6 (0.0, 16.5) |
|  | Suspect index contact | 1 | 0 | — |
| Sexual exposure to an index contact | Any index contact | 8 | 3 | 8.6 (2.1, 32.8) |
|  | Diagnosed index contact | 4 | 2 | 6.5 (1.2, 49.4) |
|  | Suspect index contact | 4 | 1 | 12.9 (2.0, 428.1) |
| Sexual exposure to an index contact with apparent symptoms at the time of the encounter | Any index contact | 1 | 2 | 1.6 (0.0, 16.5) |
|  | Diagnosed index contact | 0 | 1 | — |
|  | Suspect index contact | 1 | 1 | 3.2 (0.1, 121.0) |
| Sexual exposure to an index contact without apparent symptoms at the time of the encounter | Any index contact | 7 | 1 | 22.6 (3.8, 708.7) |
|  | Diagnosed index contact | 4 | 1 | 12.9 (1.9, 433.5) |
|  | Suspect index contact | 3 | 0 | — |

Abbreviations: OR—odds ratio; CI: confidence interval
<sup>1</sup>All analyses define individuals without any recall of exposure to an index case as the referent group (22 cases [68.8% of 32]; 71 controls [93.4% of 76]).

## Notes

### Competing Interest Statement

The authors have declared no competing interest.

### Author Declarations

As a routine public health surveillance activity, the study was considered exempt from human subjects review by the Committee for the Protection of Human Subjects of the State of California Health and Human Services Agency (project number 2022-201).

